# Mobile outreach testing for COVID-19 in twenty homeless shelters in Toronto, Canada

**DOI:** 10.1101/2020.11.23.20235465

**Authors:** Tara Kiran, Amy Craig-Neil, Paul Das, Joel Lockwood, Ri Wang, Nikki Nathanielsz, Esther Rosenthal, Carolyn Snider, Stephen W. Hwang

## Abstract

**Background:** It is unclear what the best strategy is for detecting COVID-19 among homeless shelter residents and what individual factors are associated with positivity.

**Methods:** We conducted a retrospective chart audit obtaining repeated cross-sectional data from outreach testing done at homeless shelters between April 1^st^ and July 31^st^, 2020 in Toronto, Canada. We compared the positivity rate for shelters tested because of an outbreak (at least one known case) versus surveillance (no known cases). A patient-level analysis examined differences in demographic, health, and behavioural characteristics of residents who did and did not test positive for COVID-19.

**Findings:** One thousand nasopharyngeal swabs were done on 872 unique residents at 20 shelter locations. Among the 504 tests done in outbreak settings, 69 (14%) were positive and 1 (0.2%) was indeterminate. Among the 496 tests done for surveillance, 11 (2%) were positive and none were indeterminate. Shelter residents who tested positive were significantly less likely to have a health insurance card (54% vs 72%, p=0.03) or have visited another shelter in the last 14 days (0% vs 18%, p<0.01) compared to those who tested negative; There was no association between COVID-19 positivity and medical history (p=0.40) or symptoms (p=0.43).

**Interpretation:** Our findings support testing of asymptomatic shelter residents for COVID-19 when a positive case is identified at the same shelter but suggest limited utility of testing all shelter residents in the absence of a known case. Visiting another shelter in the last 14 days is associated with a decreased risk of COVID-19 positivity.

## Introduction

On any given night, more than half a million people in the USA^1^ and more than 35,000 in Canada^2^ are homeless. Homelessness has always been associated with poor health outcomes^3^ but these risks to health have only been heightened by COVID-19, with consequences extending to the broader community. People experiencing homelessness are at increased risk of acquiring COVID-19 and spreading it to others.^4,5^ Many people experiencing homelessness stay in congregate living settings such as shelters where it is difficult to practice social distancing. Others live rough, on the street or in encampments, and do not have access to basic hygiene supplies or showering facilities. In any of these scenarios, self-isolation is not possible. People who experience homelessness also have higher rates of chronic conditions such as diabetes and hypertension which puts them at increased risk of complications if they acquire the infection. ^3,6^ Recent data from Boston suggest people experiencing homelessness have a higher prevalence of COVID-19^7^ and more severe disease^8^ than people who are housed. Modeling studies have predicted substantial near-term mortality among the homeless population in the US due to COVID-19.^9^

Strategies for testing and follow-up care impact potential spread of the virus among people experiencing homelessness.^5,10^ Outreach testing early in the pandemic confirmed high rates of COVID-19 among people experiencing homelessness who were asymptomatic but residing in a shelter with a known case.^7^ However, positivity rates have been variable when testing was done in the absence of a known case. ^11,12^ It is still unclear what the best strategy is for detecting COVID-19 among people who are homeless who live in a congregate setting, especially as community case counts decrease. It is also unclear whether, among people experiencing homelessness, there are individual demographic or health characteristics associated with acquiring COVID-19.

Our institution began conducting mobile outreach testing for COVID-19 in homeless shelters in Toronto, Canada’s largest city, approximately one month after the World Health Organization declared the global pandemic. We describe the results of mobile outreach testing at 20 different shelter locations over a three-month period. We conducted a descriptive comparison of positivity rates among shelters tested because of an outbreak (i.e. at least one known COVID-19 case at the shelter) or for surveillance (i.e. no known COVID-19 case). We also sought to assess whether there was any association between individual-level demographic, health or behavioural characteristics and COVID-19 positivity.

## Methods

### Context and Setting

Toronto is Canada’s largest city, with a total population of 2,956,024 in 2018.^13^ Toronto’s homeless population was estimated at 8,715 in 2018 with approximately 80% living in the city’s 75 shelter sites,^14^ most located in the downtown core.

St. Michael’s Hospital is located in Toronto’s downtown core. The hospital has a history of serving people experiencing homelessness and has developed partnerships to provide care at many of the nearby shelters. All permanent residents of Ontario have health insurance via the Ontario Health Insurance Plan (OHIP). Medically-necessary physician visits, hospital services, and laboratory tests are fully covered and free at the point-of-care including testing for COVID-19. During COVID-19, non-permanent residents without OHIP also have free access to testing for COVID-19.

On March 16, St. Michael’s Hospital opened one of Ontario’s COVID-19 Assessment Centres (CACs). These centres were established at locations across the province to facilitate quick and easy access to free testing for COVID-19. Testing criteria has evolved over time with an increase in testing supplies and laboratory capacity. Initially, testing criteria were restrictive. However, beginning March 18, testing was made available to symptomatic individuals who either had an occupation within an at-risk setting or were a resident in a congregate setting such as a homeless shelter. Testing criteria were slowly expanded and by June 2, testing was available to any individual, regardless of contact history, symptoms, or living situation.

Early in the pandemic, there were concerns about transmission and spread of COVID-19 in homeless shelters. In response, the St. Michael’s Hospital CAC team in partnership with Sherbourne Health began conducting mobile outreach testing to shelters in the downtown core. Initially, the CAC conducted testing in shelters on request from the local public health unit because of an outbreak situation. Outbreaks were defined as one or more cases of COVID-19 in a congregate living facility. As community case counts decreased, the focus shifted to identifying high risk settings with asymptomatic transmission. As a result, the health region began coordinating surveillance testing in shelters and directed CACs in the region to perform testing in specific shelters regardless of whether there was an outbreak. In all cases, testing was optional for shelter residents. Shelter residents, either independently or on the advice of shelter staff or medical professionals, could also choose to visit a CAC site or emergency department for testing. During this time, staff also supported shelter residents to move into hotels that were rented and repurposed by the city in an effort to reduce crowding and the related risk of COVID-19 transmission.^15^ Mobile outreach testing was also conducted at some of these hotel sites.

### Study Design and population

We conducted a retrospective audit of records from all shelter residents tested for COVID-19 by the St. Michael’s Hospital CAC mobile outreach team. Mobile outreach testing was done between April 1^st^ and July 31^st^, 2020 (further mobile testing was paused in August 2020). We excluded test results from shelter staff. We chose to focus on mobile outreach testing results and did not include charts from the main CAC site. COVID-19 testing was done using nasopharyngeal swabs performed by a physician, nurse practitioner, or registered nurse. The study was reviewed and approved by the Unity Health Toronto Research Ethics Board.

### Data collection

Age, sex, and health insurance number were collected by the outreach team at registration for all shelter residents who were tested. In some cases, the health insurance number was not available either because the resident did not have provincial health insurance (e.g. undocumented resident) or did not have the card (e.g. card lost or misplaced) and the information was not available in the hospital registration system. When sufficient staff were available for outreach, the CAC mobile outreach team also collected more detailed patient information on a standardized paper form (see Appendix) which was later scanned into the electronic medical records at St. Michael’s Hospital. The form was completed manually by trained screeners (registered nurses) at the time of testing. Screeners asked patients questions related to race, symptoms, past medical history, and shelter use in the past 14 days. The CAC team documented all COVID-19 test results in an electronic spreadsheet. Information was collected from CAC leadership on the number of people eligible for testing at a given shelter and whether testing was for outbreak or surveillance.

A team of four trained research staff reviewed twenty initial charts and compared results to ensure consistency with data extraction. The data from the rest of the charts was extracted by one of three research staff. Any uncertainties in the charts were reviewed with another member of the team and consensus was reached.

### Analysis

We performed a shelter-level descriptive analysis assessing the number of shelter residents eligible for testing and the number tested as well as the testing date and results. We categorized shelters into outbreak and surveillance depending on the purpose for outreach testing. We calculated the shelter positivity rate and plotted this over time in relation to local COVID-19 case counts.

We also performed a patient-level analysis and assessed the demographic, health, and behavioural characteristics of the shelter residents who were tested. For the subset of shelters where there was more than one COVID-19 positive resident, we compared the characteristics of residents who did and did not test positive. For residents who were tested more than once, we categorized them as testing positive if any of their results came back positive. For residents who filled more than one demographic sheet, we used the demographic responses associated with the positive test or the earliest collected non-missing response.

We also performed a patient-level descriptive analysis and assessed the demographic, health, and behavioural characteristics of the shelter residents who were tested. For residents who filled more than one demographic sheet, we used the demographic responses associated with the positive test or, if there were no positive tests or the response was missing, the earliest collected non-missing response.

For the subset of shelters where there was more than one COVID-19 positive resident, we used the same patient-level data to compare the characteristics of residents who did and did not test positive. A resident was categorized as testing positive if any of their results came back positive. We used a fixed effect logistic regression model to test whether differences in characteristics were statistically significant after adjustment for the shelter location. The overall p-value for each characteristic was calculated using a likelihood ratio test comparing a model with and without the characteristic, both adjusting for shelter location. We decided not to perform further multivariable regression analyses because of the small number of participants where we had information on health and behavioural characteristics. Chart audit data was collected in Microsoft Access and analyses were done in R version 4.0.

## Results

We conducted mobile outreach testing at 20 unique shelter locations on 25 different dates; 4 shelters were tested more than once (Table 1). Testing was conducted for a suspected outbreak between April 23 and June 1, 2020 for 430 individuals at 6 shelters on 10 different dates; between 40 and 94 percent of those living at the shelter agreed to be tested. Surveillance testing was done between June 9 and July 23, 2020 for 442 individuals at 17 shelters over 15 dates with 3 of these being hotel-sites; between 15 and 86 percent of those living at the shelter agreed to be tested. Most shelters tested for a suspected outbreak served men only while shelters tested for surveillance were more varied in the population served.

**Table 1.** Description of shelters included in mobile outreach testing.

| Shelter | Population | Testing date | Outbreak or surveillance | Number of people eligible for testing (by date) | Proportion of those eligible who were tested (by date) | Age of residents tested, Mean (SD) | Sex of residents tested, N (%) female | Residents with provincial health insurance card, N (%) |
| --- | --- | --- | --- | --- | --- | --- | --- | --- |
| Shelter A | Men | April 23 | Outbreak | 61 | 44 (72%) | 51.40 (16.24) | 0 (0.00%) | 35 (79.55%) |
| Shelter B | Mixed adults | April 30 | Outbreak | 45 | 18 (40%) | 43.92 (13.45) | 0 (0.00%) | 9 (50.00%) |
| Shelter A | Men | May 6 | Outbreak | 87 | 75 (86%) | 57.74 (10.69) | 0 (0.00%) | 71 (94.67%) |
| Shelter A | Men | May 8 | Outbreak | 57 | 34 (60%) | 45.24 (12.30) | 0 (0.00%) | 29 (85.29%) |
| Shelter C | Men | May 11 | Outbreak | 64 | 58 (91%) | 51.00 (14.19) | 3 (5.17%) | 51 (87.93%) |
| Shelter D | Mixed adults | May 20 | Outbreak | 17 | 16 (94%) | 56.04 (9.76) | 0 (0.00%) | 14 (87.50%) |
| Shelter E | Men | May 21 | Outbreak | 45 | 40 (89%) | 45.46 (11.32) | 0 (0.00%) | 17 (42.50%) |
| Shelter A | Men | May 26 | Outbreak | 87 | 75 (86%) | 57.81 (11.94) | 0 (0.00%) | 73 (97.33%) |
| Shelter F | Men | May 28 | Outbreak | 104 | 51 (49%) | 54.13 (13.78) | 0 (0.00%) | 29 (56.86%) |
| Shelter F | Men | June 1 | Outbreak | 103 | 93 (90%) | 48.85 (11.39) | 0 (0.00%) | 60 (64.52%) |
| Shelter G | Women | June 9 | Surveillance | 46 | 36 (78%) | 42.18 (11.78) | 35 (97.22%) | 21 (58.33%) |
| Shelter H | Women and Children | June 11 | Surveillance | 96 | 66 (69%) | 18.08 (15.21) | 41 (62.12%) | 32 (48.48%) |
| Shelter A | Men | June 15 | Surveillance | 81 | 56 (69%) | 43.59 (12.08) | 0 (0.00%) | 52 (89.66%) |
| Shelter I | Mixed adults | June 22 | Surveillance | 45 | 26 (58%) | 46.74 (10.37) | 2 (7.69%) | 11 (42.31%) |
| Shelter B | Mixed adults | June 24 | Surveillance | 78 | 54 (69%) | 43.35 (12.50) | 3 (5.56%) | 31 (57.41%) |
| *Shelter J | Men | June 29 | Surveillance | 47 | 35 (74%) | 43.94 (13.17) | 0 (0.00%) | 24 (68.57%) |
| Shelter C | Men | July 2 | Surveillance | 64 | 55 (86%) | 48.15 (14.07) | 0 (0.00%) | 48 (88.89%) |
| Shelter K | Youth | July 6 | Surveillance | 100 | 21 (21%) | 22.09 (3.31) | 4 (22.22%) | 9 (42.86%) |
| Shelter L | Women | July 7 | Surveillance | 24 | 12 (50%) | 52.50 (13.41) | 10 (83.33%) | 11 (91.67%) |
| Shelter M | Women | July 7 | Surveillance | 25 | 16 (64%) | 65.47 (9.64) | 16 (100.00%) | 15 (93.75%) |
| Shelter N | Women | July 8 | Surveillance | 40 | 15 (38%) | 48.93 (16.70) | 15 (100.00%) | 1 (6.67%) |
| Shelter O | Mixed Adults | July 8 | Surveillance | 40 | 18 (45%) | 50.29 (14.56) | 0 (0.00%) | 9 (50.00%) |
| Shelter P | Mixed Adults | July 14 | Surveillance | 40 | 14 (35%) | 50.13 (11.52) | 3 (21.43%) | 10 (71.43%) |
| Shelter Q | Female | July 15 | Surveillance | 33 | 10 (30%) | 41.46 (11.46) | 10 (100.00%) | 3 (30.00%) |
| *Shelter R | Mixed Adults | July 21 | Surveillance | 197 | 30 (15%) | 52.63 (12.91) | 9 (30.00%) | 16 (53.33%) |
| Shelter S | Youth | July 22 | Surveillance | 9 | 6 (67%) | 23.58 (1.77) | 0 (0.00%) | 4 (66.67%) |
| *Shelter T | Young Mixed Adults | July 23 | Surveillance | 65 | 25 (38%) | 37.71 (9.37) | 8 (32.00%) | 20 (80.00%) |

\*Hotel site

The outreach team conducted 1000 tests for 872 unique shelter residents (504 tests in outbreak settings and 496 tests for surveillance). The demographic and health characteristics of unique shelter residents tested are summarized in Table 2. The mean age was 46, 82% were men, and 68% were able to provide a valid provincial health insurance card. More detailed demographic information was available for a subset of 348 individuals: 40% were white, 52% were racialized, and 8% chose not to disclose their racial identity. 82% reported a health condition with approximately one-quarter (27%) saying they had a mental health condition, almost a third (31%) disclosing a substance use disorder, and over half (55%) stating they smoked cigarettes. Only 9% reported any symptoms of COVID-19 with the most common symptom being a cough. 8% reported visiting another shelter in the last 14 days.

**Table 2.** Sociodemographic and health characteristics, shelter use and symptom prevalence for unique individuals tested during mobile outreach

|  | All [N (%)] |
| --- | --- |
| Total | 872 |
| Age, N=871 |  |
| 0-15 | 38 (4.36%) |
| 16-24 | 37 (4.25%) |
| 25-49 | 417 (47.88%) |
| 50-64 | 277 (31.80%) |
| 65+ | 102 (11.71%) |
| Age, mean (SD) | 45.81 (16.31) |
| Sex, N=868 |  |
| N (%) Female | 159 (18.32%) |
| Provincial health insurance number available, N=872 | 589 (67.55%) |
| Race*, N=335 |  |
| Black | 77 (22.99%) |
| East/Southeast Asian | 23 (6.87%) |
| Latino | 9 (2.69%) |
| Middle Eastern | 9 (2.69%) |
| South Asian | 22 (6.57%) |
| White | 134 (40.00%) |
| Other (includes indigenous) | 34 (10.15%) |
| Prefer not to disclose | 27 (8.06%) |
| Past Medical History*, N=304 |  |
| Any chronic condition | 248 (81.58%) |
| CV disease | 17 (5.59%) |
| Chronic lung disease | 19 (6.25%) |
| HIV | 9 (2.96%) |
| Diabetes | 26 (8.55%) |
| Current smoker | 168 (55.26%) |
| Mental health diagnosis | 82 (26.97%) |
| Substance use | 93 (30.59%) |
| Other | 38 (12.50%) |
| Prefer not to disclose | 25 (8.22%) |
| Symptoms*, N=331 |  |
| Any symptoms | 32 (9.20%) |
| Cough | 20 (6.04%) |
| Shortness of Breath | <2% |
| Fever | 0 (0.00%) |
| Other | 12 (3.45%) |
| Visited another shelter in last 14 days*, N=348 | 41 (11.78%) |

\* The race, past medical history, symptoms and visited another shelter variables based on demographic info collected for only 348 respondents

Among the 504 tests done in outbreak settings, 69 (14%) were positive and 1 (0.2%) was indeterminate. In subsequent analysis, we excluded the indeterminate result. Among the 496 tests done for surveillance, 11 (2%) were positive and none were indeterminate.

Figure 1 summarizes the positivity rate by shelter and testing date in relation to the total new number of COVID-19 cases in Toronto. Among the 10 testing dates done because of an outbreak, 2 found no positive cases and 1 had a single positive case; the positivity rate for the remaining dates ranged from 4% to 33%. Only 1 of 17 shelters tested for surveillance had any positive cases. Outbreak testing and related positivity occurred between April and early June, when case counts in Toronto were highest. In 6 of 10 instances of outbreak testing, the positivity rate was the same or higher than the average positivity rate in Toronto (Appendix).

**Figure 1:**
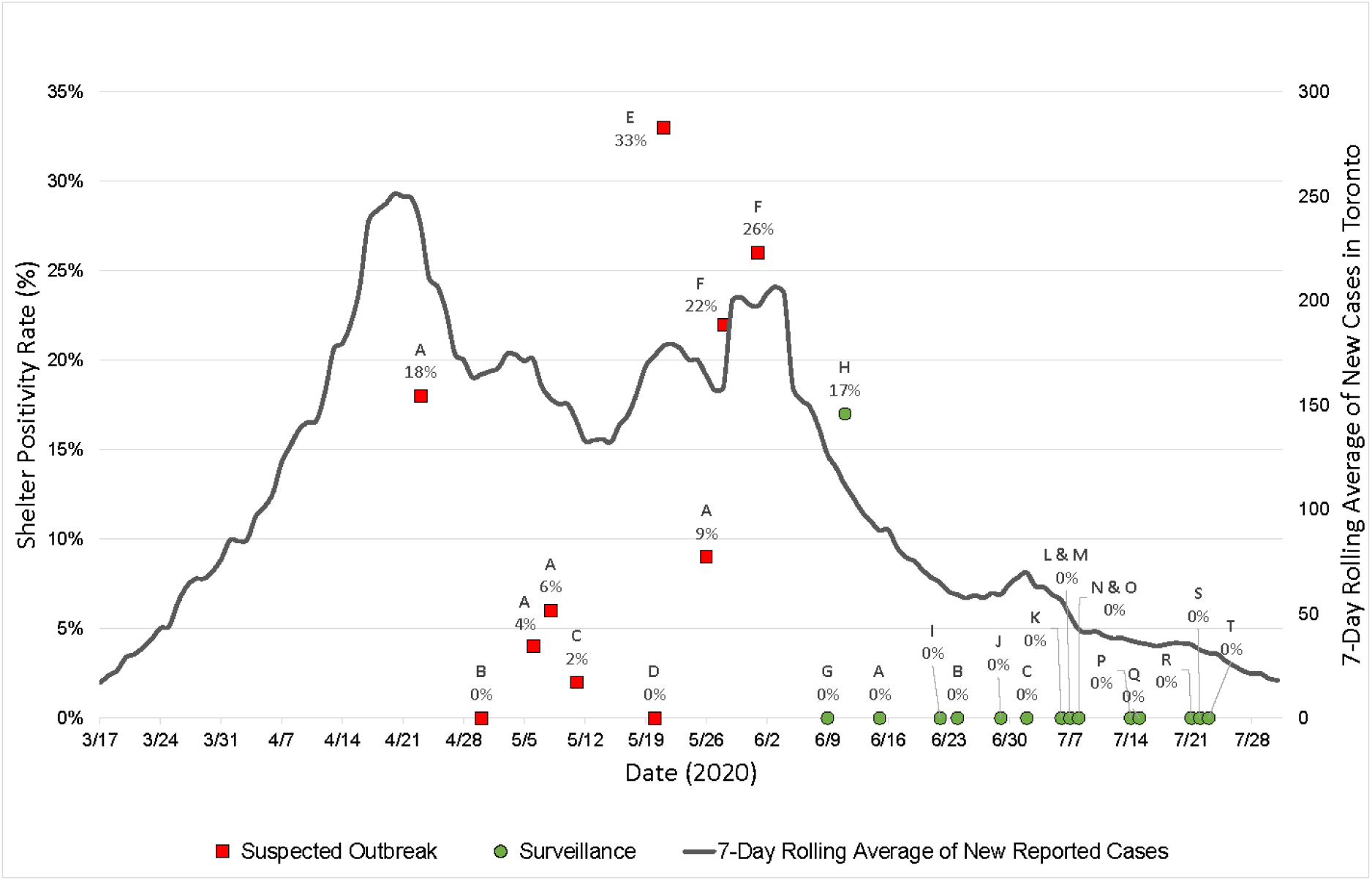
Shelter Positivity Rate vs 7-Day Rolling Average of New COVID-19 Cases in Toronto.

We compared the demographic characteristics between shelter residents who tested positive and negative for COVID-19. We included data from the 8 testing dates at four 4 unique shelter locations where more than one individual tested positive (Table 3). Residents who tested positive were less significantly less likely to have a provincial health insurance card (54% vs 72%, p=0.03) and significantly less likely to have visited another shelter in the last 14 days (0% vs 18%, p<0.01) compared to those who tested negative. Residents who tested positive also had a higher mean age compared to those who tested negative (48.3 vs 45.5, p=0.08) and a higher proportion were racialized (62% vs 48%, p=0.20), but these differences were not statistically significant. There were no differences between groups related to presence of any medical condition or any symptom.

**Table 3.** Comparison of sociodemographic and health characteristics, symptom prevalence, and shelter use for individuals who did and did not test positive for COVID-19 at shelter test dates where there was more than one positive case. The following shelters and dates were included: Shelter A – April 23 Shelter A – May 6 Shelter A – May 8 Shelter E – May 21 Shelter A – May 26 Shelter F – May 28 Shelter F – June 1 Shelter H – June 11

|  | Positive for<br>COVID-19<br>N (%) | Negative for<br>COVID-19<br>N (%) | P-value |
| --- | --- | --- | --- |
| All | 78 (19.31%) | 326 (80.69%) |  |
| Age |  |  |  |
| 0-15 | 7 (8.97%) | 31 (9.51%) |  |
| 16-49 | 34 (43.59%) | 141 (43.25%) |  |
| 50-64 | 23 (29.49%) | 113 (34.66%) |  |
| 65+ | 14 (17.95%) | 41 (12.58%) | 0.31 |
| Age, mean (SD) | 48.32 (17.99) | 45.46 (18.47) | 0.08 |
| Sex |  |  |  |
| Female | 6 (7.69%) | 35 (10.74%) | 0.57 |
| Provincial health insurance number<br>available | 42 (53.85%) | 236 (72.39%) | 0.03 |
| Race* |  |  |  |
| No. of individuals with responses | 42 | 189 |  |
| Racialized | 26 (61.90%) | 90 (47.62%) |  |
| White | 12 (28.57%) | 82 (43.49%) |  |
| Prefer not to disclose | 4 (9.53%) | 17 (8.99%) | 0.20 |
| Past Medical History* |  |  |  |
| No. of individuals with responses | 36 | 173 |  |
| Any chronic condition | 28 (77.78%) | 142 (82.80%) | 0.40 |
| Symptoms* |  |  |  |
| No. of individuals with responses | 44 | 198 |  |
| Any symptom | 5 (11.36%) | 17 (8.59%) | 0.43 |
| Visited another shelter in last 14 days* |  |  |  |
| No. of individuals with responses Visited<br>another shelter | 44<br>0 (0.00%) | 198<br>35 (17.68%) | <0.01 |

\* The race, past medical history, symptoms and visited another shelter variables based on demographic info collected for only 44 respondents in the positive category and 198 in the negative category.

### Interpretation

We conducted 1000 tests for SARS-CoV-2 through mobile outreach testing at twenty shelter locations in Canada’s largest city between April 23 and July 23, 2020, during which time the number of new daily COVID-19 cases in the city dropped from 237 to 31. Approximately half of the tests were done because of a suspected outbreak and half for surveillance, with the former coinciding with higher numbers of new cases in the city. We found that 14% of tests done in an outbreak setting were positive compared to 2% done for surveillance. We found no association between COVID-19 positivity and the presence of any medical history or any symptoms. Shelter residents who tested positive were significantly less likely to have a provincial health insurance card or visit another shelter in the last 14 days. Our analysis also suggested that shelter residents who tested positive were more likely to be older in age and identify as racialized but these differences were not statistically significant.

People experiencing homelessness are known to be vulnerable to COVID-19^4,5^ and our study confirms this as we found high rates of positivity in shelter residents relative to the general population. However, even within the shelter population, we found different degrees of vulnerability. Those who visited another shelter in the previous two weeks were less likely to test positive perhaps because they were spending more time outdoors where there is thought to be a lower risk of transmission of SARS-CoV-2.^16^ In our setting, shelter residents could also have received testing in the emergency department or a COVID-19 Assessment Centre. Residents without a provincial health insurance card were more likely to test positive perhaps because they delayed seeking testing for COVID-19 or because they faced more barriers to protecting themselves from infection such as language, income, or more chaotic life circumstances. Racialized groups have been harder hit by COVID-19 in Canada^17^ and the US.^18,19^ Our data suggests that homelessness and race may be intersecting factors that increase vulnerability and supports calls to address issues of structural racism at the root of poor outcomes.^20^

Only a handful of other studies have reported on testing for SARS-CoV-2 in homeless shelters, most in the US. Mosites and colleagues describe low positivity rates among shelter residents undergoing surveillance testing in Atlanta but note positivity rates of 17%, 36%, and 66% among residents included in outbreak testing done in Seattle, Boston, and San Francisco, respectively.^11^ Consistent with our findings, researchers in Boston found almost 90% of residents who tested positive in the context of an outbreak were asymptomatic.^7^ Surveillance screening at five shelters in Rhode Island found an overall positivity rate of 12%--higher than in our setting--but like in our study, symptom prevalence did not vary between those who did and did not test positive.^12^ The only other Canadian study, from Hamilton, Ontario, used a strategy of testing shelter residents who screened positive for symptoms and reported a very low positivity rate;^21^ however overall case counts have been much lower in Hamilton compared to Toronto.

Our findings support testing of asymptomatic individuals living at a shelter where another individual has tested positive for SARS-CoV-2. However, our results call into question the utility of surveillance testing in shelters with no known positive cases, particularly when community case counts are low. We found only one of seventeen shelters tested for surveillance in our setting had any positive cases. This shelter had been the site of an outbreak 3 weeks prior and was also the only family shelter tested. Children are known to have milder disease and many studies have found asymptomatic COVID-19 infection in children.^22^ Together, these findings suggest that surveillance testing of all shelter residents is of limited value when community case counts are low with the possible exception of congregate settings with children. Sentinel surveillance testing, ^23,24^ through nasopharangeal swabs or saliva sampling,^25^ are other strategies that warrant further study.

Our study has several limitations. First, our study is based on testing done by our institution’s mobile outreach team in shelters over three months. A more comprehensive picture would include results from individuals experiencing homelessness who were tested in the emergency department, COVID-19 Assessment Centres, and by other mobile outreach teams and include individuals sleeping rough. Second, testing was voluntary for shelter residents which may introduce selection bias and influence positivity rates. Third, our ability to detect statistically significant differences between those who did and did not test positive was limited by our sample size and the number of demographic questionnaires completed in our sample. Fourth, the difference in positivity rates from outbreak and surveillance may be explained by the difference in community case counts and the changes to shelters to reduce transmission over the same time period. Finally, we did not engage people with lived experience of homelessness as research partners but hope to do so in future work.

In summary, we found a COVID-19 positivity rate of 14% among residents of homeless shelters tested because of a known outbreak (i.e. at least one known case) and 2% among residents when testing was done for surveillance (i.e. no known cases). There was no difference in health history or symptoms between residents who tested positive and negative but there were differences related to health insurance status, age, race, and use of other shelters. Our findings support testing of asymptomatic shelter residents for COVID-19 when a positive case is identified at the same shelter but suggest limited utility of surveillance testing of all shelter residents with the possible exception of shelters that include children. Our results suggest that when homelessness intersects with factors such as race and health insurance status, individuals experience increased vulnerability to COVID-19 infection. Research and policy interventions should seek to understand and address these individual-level factors. Ultimately, solutions to the increased risk of COVID-19 transmission among people experiencing homeless need to address root causes of homelessness including affordable housing, a living wage, and adequate social supports.

## Supporting information

STROBE statement

## Data Availability

Due to the nature of this research, participants of this study did not agree for their data to be shared publicly, so supporting data is not available.

## Acknowledgements

We are grateful to Linh Luong and Tadios Tibebu who spent many hours extracting data from paper charts. We thank the many individuals involved in supporting the mobile outreach testing, particularly Dana Whitman, Nicole Gichuru, Chantel Marshall and Linda Jackson, whose leadership and guidance made the outreach possible. We would like to especially acknowledge the daily hard work by staff at the COVID-19 Assessment Centre at St. Michael’s Hospital, Sherbourne Health, and the shelter sites.

