## Appendices for "Mobile outreach testing for COVID-19 in twenty homeless shelters in Toronto, Canada"

**Appendix**

**Exhibit 1: Sample CAC data collection form**

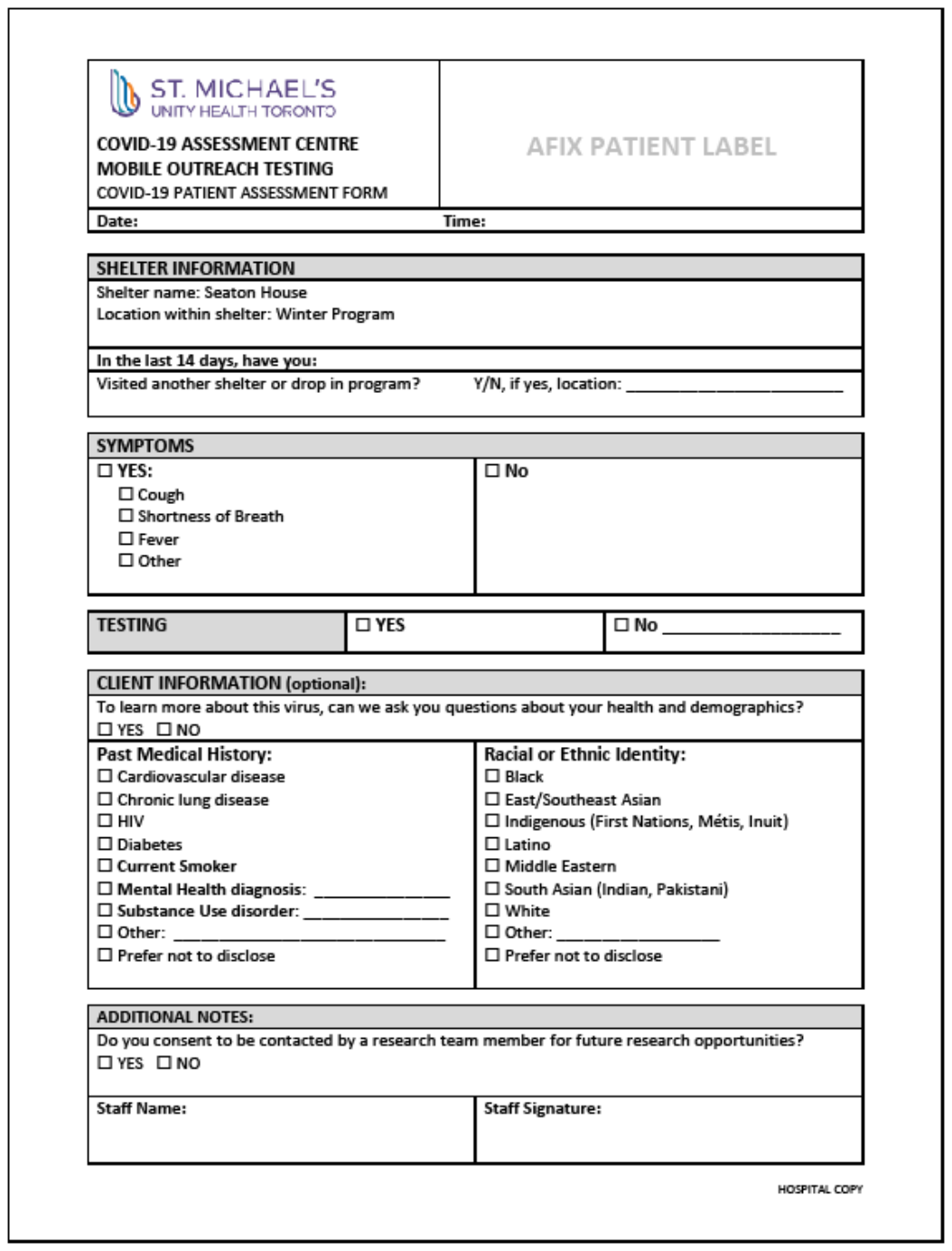

**Exhibit 2: Shelter Positivity Rate vs. Weekly Positivity Rate in Toronto**

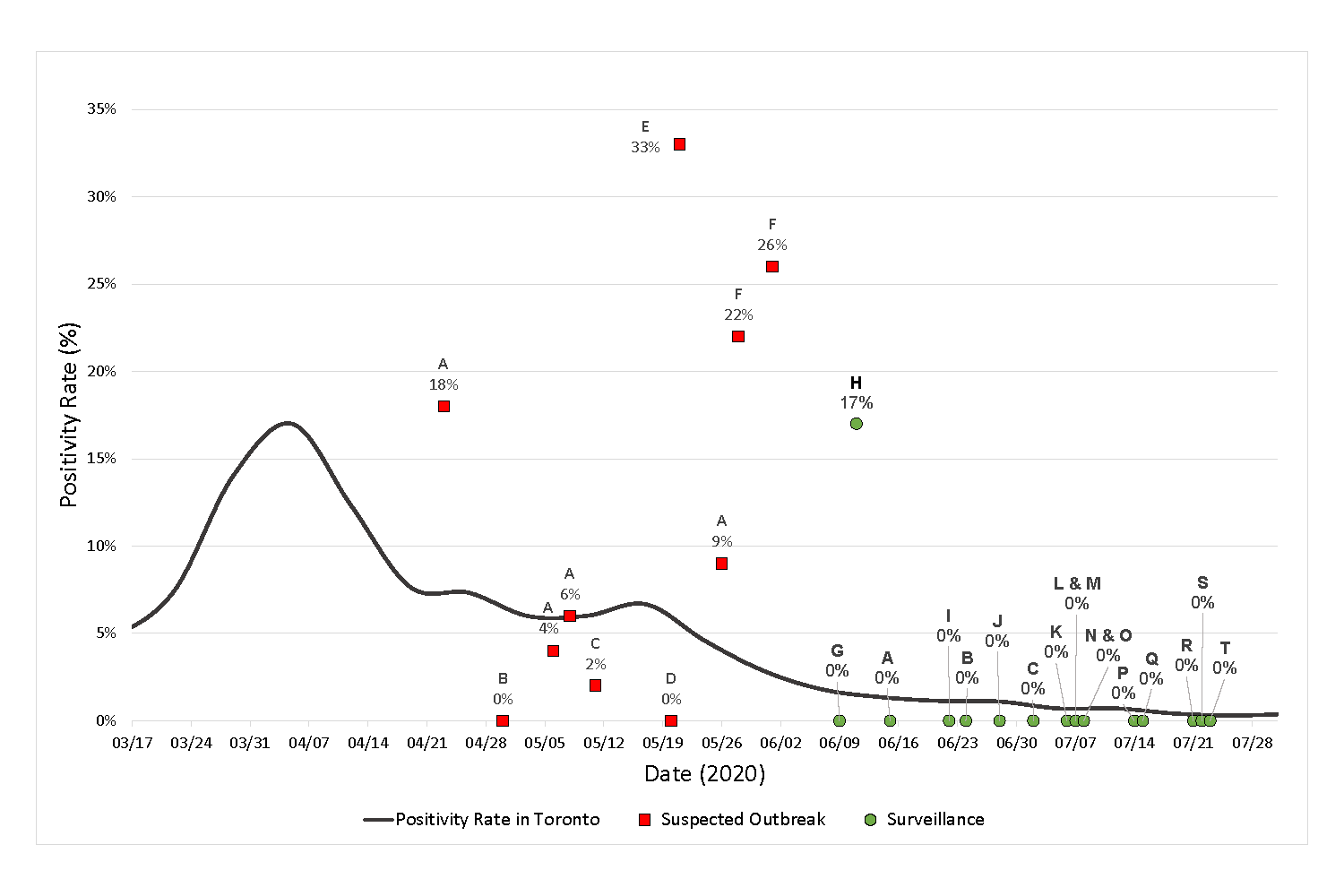
